# Severity of respiratory failure and computed chest tomography in acute COVID-19 correlates with pulmonary function and respiratory symptoms after infection with SARS-CoV-2: an observational longitudinal study over 12 months

**DOI:** 10.1101/2021.08.11.21261883

**Authors:** Fridolin Steinbeis, Charlotte Thibeault, Felix Doellinger, Raphaela Maria Ring, Mirja Mittermaier, Christoph Ruwwe-Glösenkamp, Florian Alius, Philipp Knape, Hans-Jakob Meyer, Lena Johanna Lippert, Elisa Theresa Helbig, Daniel Grund, Bettina Temmesfeld-Wollbrück, Norbert Suttorp, Leif Erik Sander, Florian Kurth, Tobias Penzkofer, Martin Witzenrath, Thomas Zoller

## Abstract

**Background:** Prospective and longitudinal data on pulmonary injury over one year after acute coronavirus disease 2019 (COVID-19) are sparse.

**Research question:** With this study, we aim to investigate pulmonary outcome following SARS-CoV-2 infection including pulmonary function, computed chest tomography, respiratory symptoms and quality of life over 12 months.

**Study design and Methods:** 180 patients after acute COVID-19 were enrolled into a single-centre, prospective observational study and examined 6 weeks, 3, 6 and 12 months after onset of COVID-19 symptoms. Chest CT-scans, pulmonary function and symptoms assessed by St. Georges Respiratory Questionnaire were used to evaluate objective and subjective respiratory limitations. Patients were stratified according to acute COVID-19 disease severity.

**Results:** Of 180 patients enrolled, 42/180 were not hospitalized during acute SARS-CoV-2 infection, 29/180 were hospitalized without need for oxygen, 43/180 with need for low-flow and 24/180 with high-flow oxygen, 26/180 required invasive mechanical ventilation and 16/180 were treated with ECMO. After acute COVID-19, pulmonary restriction and reduced carbon monoxide diffusion capacity was associated with disease severity after the acute phase and improved over 12 months except for those requiring ECMO treatment. Patients with milder disease showed a predominant reduction of ventilated area instead of simple restriction. The CT score of lung involvement in the acute phase increased significantly with COVID-19 severity and was associated with restriction and reduction in diffusion capacity in follow-up. Respiratory symptoms improved for patients in higher severity groups during follow-up, but not for patients with mild initially disease.

**Interpretation:** Severity of respiratory failure during COVID-19 correlates with the degree of pulmonary function impairment and respiratory quality of life in the year after acute infection. Patients with mild vs. severe disease show different patterns of lung involvement and symptom resolution.

**Clinical Trial Registration:** The study is registered at the German registry for clinical studies (DRKS00021688)

## Background

Severe acute respiratory syndrome corona virus 2 (SARS-CoV-2) causes acute viral respiratory tract infections including pneumonia. After initial infection with SARS-CoV-2 in the upper respiratory tract, viral replication continues in lower airways and alveolar epithelial cells ^37^, leading to a hyper-inflammatory immune response causing alveolar damage and vascular leakage ^30, 38^. Chronic lung injury was observed in 25-63% patients three months post-acute COVID-19^21, 32^. Known pathomechanisms of chronic lung injury and fibrosis such as a TGF-beta dominated adaptive immune response ^6^, fibroblast activation ^11^, alveolar epithelial cell death and distortion of the basal lamina leading to alveolar collapse induration ^26^ have been observed in COVID-19. Moreover, viral pneumonia following severe acute respiratory syndrome corona virus (SARS), Middle East Respiratory Syndrome Coronavirus (MERS) and Influenza-A-Virus H1N1 (H1N1) infections have been associated with pulmonary restriction and reduced diffusion capacity and pathological chest CT findings ^17, 14, 28^. First data of the early post-acute COVID-19 phase revealed that up to four months post infection, COVID-19 patients show a pattern of pulmonary restriction and abnormal carbon monoxide diffusion capacity in lung function testing ^3, 16, 22, 25, 40, 13, 21, 32^. Similar results were seen for month 6 and 12 after symptom onset in a Chinese prospective cohort study ^15, 39^.

So far, prospective and longitudinal data on pulmonary injury over one year after acute coronavirus disease 2019 (COVID-19) are sparse, particularly no data from European patients are available. Further, disease severity is commonly classified according to WHO groups into mild/moderate and severe and critical. With this prospective study, we aim to provide high-resolution data on pulmonary function, symptom burden, patient reported outcomes and radiological characteristics of SARS-CoV-2 infection in more detail. We stratified patients into six severity categories and observed them over a period of 12 months. Moreover, this study aims to describe different patterns of pulmonary injury and their relation to subjective limitations in hospitalized and non-hospitalized patients with COVID-19.

## Methods

### Patients

Adult patients with evidence of SARS-CoV-2 infection (determined by positive SARS-CoV-2 PCR), were enrolled in the Pa-COVID-19 study at our tertiary care centre (Charité-Universitätsmedizin Berlin, Germany) ^19^. Pa-COVID-19 is a prospective observational study collecting clinical data and biosamples during hospital treatment and at outpatient follow-up visits. Patients were included either at the time of hospital admission for acute SARS-CoV-2 infection (138; 76.7%) or during follow-up (42; 23.3%) for patients not initially included at our hospital. All participants or their legal representatives gave written informed consent before study inclusion. The study was approved by Charité ethics committee (EA2/066/20). This analysis includes patients who were followed-up as outpatients between May 2020 and June 2021. Patients were examined 6 weeks as well as 3, 6 and 12 months after onset of first symptoms of SARS-CoV-2 infection.

### COVID-19 severity groups

Patients were stratified by acute COVID-19 severity in analogy to the WHO ordinal scale of clinical improvement ^36^, into i) non-hospitalized patients without supplemental oxygen therapy (NOO), ii) hospitalized patients without supplemental oxygen therapy (NOH), iii) with supplemental low-flow oxygen therapy (LFO), iv) high-flow oxygen therapy (HFO), v) invasive mechanical ventilation (IMV) and vi) extracorporeal membrane oxygenation (ECMO). Patients were categorized according to the highest severity of respiratory failure as expressed by the level of respiratory support which occurred during acute COVID-19. Treatment allocation with regard to type of respiratory support was not limited by available medical resources during the study period, but was guided by current clinical guidelines for patients with need for ECMO.

### Pulmonary function tests

Pulmonary function was examined using Ganshorn PowerCube Body+ and Diffusion+ (Schiller Group, Niederlauer, Germany) and performed according to the German, European and American recommendations for pulmonary function testing ^29, 5, 10^. Reference values were calculated based on the Global Lung Function Initiative (GLI) reference equations (GLI-2012) and results are expressed as percent predicted value (ppv) ^33^. Interpretation and grading of diffusing capacity values was adapted from the ERS/ATS official technical standards and the subsequent correspondence ^8, 9^. Pulmonary restriction or obstruction was defined according to the “ATS/ERS Task Force: Standardisation of Lung Function Testing” as TLC <5^th^ percentile of the lower limit of normal (LLN) and FEV1/FVC < LLN ^20^. Complex restriction was defined according to Clay et al. as difference between ppv TLC and FVC >10% ^4^. No further breakdown into severity grades was performed for categorical analysis.

### Chest computed Tomography (CT)

CT-scans were performed on the basis of clinical guideline recommendations. If available, the first CT scan performed within 30 days after symptom onset was used for analysis. Chest-CT scans were reviewed by two senior thoracic radiologists. All images were reviewed blinded to the patient’s clinical characteristics and disease severity. Pulmonary involvement during the acute phase was assessed using a visual score ranging from 0 (no involvement) to 5 (>75% involvement) for each lung lobe as described in more detail by Pan et al. ^27^.

### Symptom assessment and health related quality of life

A standardized list of 43 symptoms was evaluated at each study visit at baseline and during follow-up in a patient interview (Table S3). To capture overall impact on health, daily life and wellbeing in patients, the St. George’s Respiratory Questionnaire (SGRQ) was measured^7^. A total score of 25 or higher, as suggested by the Global strategy for the diagnosis, management, and prevention of chronic obstructive pulmonary disease, was used as threshold for limitations in health and wellbeing.

### Data analysis

Descriptive statistics was used to calculate median, inter-quartile range (IQR), mean and standard deviations (SD). Difference in continuous variables between three or more groups were analysed by one-way ANOVA or Kruskal-Wallis test. Fischer’s exact test (for sample size <5 per group) or Chi-square test were used for analysis of categorical variables. The correlation between lung function and patient reported outcomes from SGRQ was calculated using Pearson correlation coefficients with a two-sided 95% confidence interval. Logistic regression was performed to assess association of clinical variables, radiological findings and patient reported outcomes with pulmonary restriction and reduced D_LCO_ in post-acute COVID-19. For univariate and multivariate analysis of risk factors for pulmonary restriction and diffusion capacity, patient characteristics and comorbidities recorded at the study inclusion visit were used and for pulmonary function and SGRQ, the lowest values observed during follow-up was used. Variables were adjusted for confounders as determined by clinical evidence (age, BMI) ^12^ or due to a significant relationship in univariate testing (i.e. gender, disease severity). Statistical significance was assumed for p<0.05. The level of significance is marked with asterisks; ^*^ for p < 0.05; ^**^ p < 0.01; ^***^ p < 0.001 and ^****^ p < 0.0001. IBM SPSS (IBM SPSS Statistics 27.0), JMP (version 14.2.0) and GraphPad PRISM (Version 9.0.0) were used for statistical analysis and graphical processing.

## Results

### Baseline characteristics

180 patients who presented to our outpatient department following acute COVID-19 with at least one complete pulmonary function dataset 6 weeks, 3-, 6-, and 12 months after symptom onset were included. A smaller number of patients presented at 6 week follow-up as many patients were either still in inpatient treatment or at rehabilitation at this point of time. At the time of analysis, 73/180 patients (40.5%) participated at follow-up visits at week 6, 118/180 (65.5%) at month 3, 139/180 (77.2%) at month 6 and 72/180 (40.0%) at month 12 of follow-up. 13/180 patients (7.2%) were lost to follow-up.

138/180 (76.6%) patients were initially hospitalized. 42/180 (23.3%) were never hospitalized (NOO), 29/180 (16.1%) were treated on a normal ward without need for oxygen (NOH), 43/180 (23.9%) received low-flow oxygen treatment via nasal cannula (LFO), 24/180 (13.3%) received high-flow oxygen treatment (HFO), 26/180 (14.4%) patients required invasive mechanical ventilation (IMV) and 16/180 (8.8%) patients were treated with ECMO (determined by the highest level of respiratory support, Table 1).

**Table 1:** Patient characteristics stratified by level of respiratory support during acute phase of COVID-19. Significant differences were shown for age, BMI, chronic heart disease, diabetes, pulmonary restriction and D_LCO_ reduction (bold). Statistical significance is calculated by Chi2, Fischer’s exact test or Kruskal-Wallis-test where applicable. Abbreviations: NOO – no oxygen, outpatient; NOH – no oxygen hospitalized; LFO – low-flow oxygen supply; HFO – high-flow oxygen supply; IMV – invasive mechanical ventilation; ECMO – extracorporeal membrane oxygenation; CCI – Charlson Comorbidity Index. ^*^missing values: Smoking history n=3; Immunosuppression: n=1.

|  | ALL<br>(n=180) | LEVEL OF RESPIRATORY SUPPORT |  |  |  |  |  |  |
| --- | --- | --- | --- | --- | --- | --- | --- | --- |
|  |  | NOO (n = 42) | NOH (n = 29) | LFO (n = 43) | HFO (n = 24) | IMV (n = 26) | ECMO (n = 16) | p |
| Age (median (IQR)) | 57 (43.25-65.75) | 44 (35-60) | 62 (40-70) | 56 (48-65) | 60 (52-66) | 61 (52-71) | 56 (49-64) | <0.0001 |
| Sex female (n / %) | 68 (37.8%) | 26 (61.9) | 15 (51.72) | 10 (23.26) | 5 (20.83) | 7 (26.92) | 5 (31.25) | 0.001 |
| Sex male (n / %) | 112 (62.2%) | 16 (38.10) | 14 (48.28) | 33 (76.74) | 19 (79.17) | 19 (65.52) | 11 (68.75) | 0.001 |
| BMI (median (IQR)) | 26.72 (23.88-31.3) | 24.02 (22.45-26.47) | 25.1 (23.46-27.34) | 27.76 (24.33-32.72) | 29.52 (25.95-33.36) | 29.39 (26.12-32.14) | 29.35 (27.56-35.51) | 0.004 |
| SMOKING |  |  |  |  |  |  |  |  |
| Smoking history* (n / %) | 62 (34.4%) | 12 (29.27%) | 8 (27.59%) | 16 (37.21%) | 8 (33.33%) | 14 (56.00%) | 4 (26.67%) | 0.246 |
| COMORBIDITIES |  |  |  |  |  |  |  |  |
| CCI 0 (n / %) | 51 (28.3%) | 21 (50.00%) | 8 (27.59%) | 10 (23.26%) | 5 (20.83%) | 3 (11.54%) | 4 (25.00%) | 0.016 |
| CCI 1-2 (n / %) | 65 (36.1%) | 13 (30.95%) | 7 (24.14%) | 20 (46.51%) | 9 (37.50%) | 10 (38.46%) | 6 (37.50%) | 0.502 |
| CCI 3-4 (n / %) | 46 (25.6%) | 8 (19.05%) | 14 (48.28%) | 8 (18.60%) | 4 (16.67%) | 8 (30.77%) | 4 (25.00%) | 0.063 |
| CCI >5 (n / %) | 18 (10%) | 0 | 0 | 5 (11.63%) | 6 (25.00%) | 5 (19.23%) | 2 (12.50%) | 0.001 |
| Chronic lung disease (n / %) | 26 (14.4%) | 6 (14.29%) | 4 (13.79%) | 2 (4.65%) | 5 (20.83%) | 6 (23.08%) | 3 (18.75%) | 0.224 |
| Asthma (n / %) | 13 (7.2%) | 5 (11.90%) | 1 (3.45%) | 1 (2.33%) | 2 (8.33%) | 4 (15.38%) | 0 | 0.205 |
| COPD (n / %) | 11 (6.1%) | 0 | 2 (6.90%) | 2 (4.65%) | 2 (8.33%) | 3 (11.54%) | 2 (12.50%) | 0.166 |
| Chronic heart disease (n / %) | 78 (43.3%) | 9 (21.43%) | 10 (34.48%) | 19 (44.19%) | 14 (58.33%) | 15 (57.69%) | 11 (68.75%) | 0.003 |
| Chronic kidney disease (n / %) | 21 (11.7%) | 2 (4.76%) | 6 (20.69%) | 6 (13.95%) | 2 (8.33%) | 3 (11.54%) | 2 (12.50%) | 0.429 |
| Diabetes (n / %) | 30 (16.7%) | 4 (9.52%) | 1 (3.45%) | 6 (13.95%) | 7 (29.17%) | 5 (19.23%) | 7 (43.75%) | 0.006 |
| Chronic liver disease (n / %) | 9 (5%) | 1 (2.38) | 2 (6.90%) | 2 (4.65%) | 2 (8.33%) | 2 (7.69%) | 0 | 0.777 |
| Chronic immunological disease (n / %) | 4 (5.3) | 2 (4.76) | 4 (13.79) | 2 (4.65) | 2 (8.33) | 0 | 3 (18.75%) | 0.139 |
| Chronic neurological disease (n / %) | 23 (12.8%) | 4 (9.52%) | 3 (10.34%) | 8 (18.60%) | 5 (20.83%) | 1 (3.85%) | 2 (12.50%) | 0.405 |
| Psychiatric disease (n / %) | 12 (6.7%) | 3 (7.14%) | 1 (3.45%) | 3 (6.98%) | 2 (8.33%) | 0 | 3 (18.75) | 0.290 |
| Active cancer (n / %) | 5 (2.8%) | 2 (4.76%) | 1 (3.45%) | 0 | 0 | 0 | 0 | 0.756 |
| Chronic haematological disease (n / %) | 4 (2.2%) | 1 (2.38%) | 2 (7.14%) | 3 (6.98%) | 4 (16.67%) | 0 | 1 (6.25%) | 0.188 |
| Immunosuppression* (3M pre-COVID) (n / %) | 11 (6.1%) | 1 (2.38%) | 2 (7.14%) | 3 (6.98%) | 4 (16.67%) | 0 | 1 (6.25%) | 0.188 |
| Organ transplanted (n / %) | 5 (2.8%) | 1 (2.38%) | 2 (6.90%) | 1 (2.33%) | 1 (4.17%) | 0 | 0 | 0.779 |
| PULMONARY FUNCTION |  |  |  |  |  |  |  |  |
| Simple Restriction (n / %) | 59 (32.8%) | 5 (11.90%) | 5 (17.24%) | 13 (30.95%) | 11 (45.83%) | 16 (61.54%) | 9 (56.25%) | <0.0001 |
| Complex Restriction (n / %) | 92 (51.10%) | 25 (59.52%) | 19 (65.52%) | 19 (45.24%) | 12 (50.00%) | 10 (38.46%) | 7 (43.75%) | 0.337 |
| Obstruction (n / %) | 8 (4.40%) | 0 | 1 (3.45%) | 2 (4.76%) | 2 (8.33%) | 3 (11.53%) | 0 | 0.185 |
| D<sub>LCO</sub> reduced (n / %) | 109 (60.60%) | 18 (42.86%) | 15 (51.72%) | 26 (61.90%) | 26 (61.90%) | 17 (65.38%) | 19 (76.00%) | 0.010 |

Median age of all patients was 56.5 years (IQR 43.25-65.75). Median age of patients increased continuously with the level of respiratory support from 44 years (35-60) in the NOO group to 61 years (52-71) in the MV group, mean age in the ECMO group was 56 years (49-64) (Table 1). Median (IQR) body-mass index (BMI) of study participants increased with level of respiratory support. Overall, 68/180 (37.8%) of all study participants were female, and the proportion of female patients was reduced with increasing level of care. Of all patients, 62/180 (34.4%) were former or current smokers. COVID-19 severity correlated particularly with chronic heart disease and diabetes (Table 1).

### Body plethysmography, carbon monoxide diffusion capacity and respiratory muscle strength

55/180 (32%) study participants showed pulmonary restriction and 104/180 (61%) showed reduced carbon monoxide diffusion capacity (D_LCO_) (lowest value at any point of time during follow-up). Pulmonary restriction and impairment of D_LCO_ was significantly associated with increasing severity of lung failure expressed as the level of respiratory support during the acute phase of SARS-CoV-2 infection (Table 1). In contrast, complex restriction, as defined by Clay et al. ^4^, was not associated with severity of respiratory failure during acute SARS-CoV-2 infection, but was slightly more common among patients with mild disease.

Significant differences in restrictive ventilation patterns were seen between groups of different disease severity during acute COVID-19. (Figure 1, Table s1). Median (IQR) of TLC and FVC was significantly lower in patients with higher level of respiratory support during acute COVID-19. This difference persisted until 12-months after acute COVID-19.

**Figure 1:**
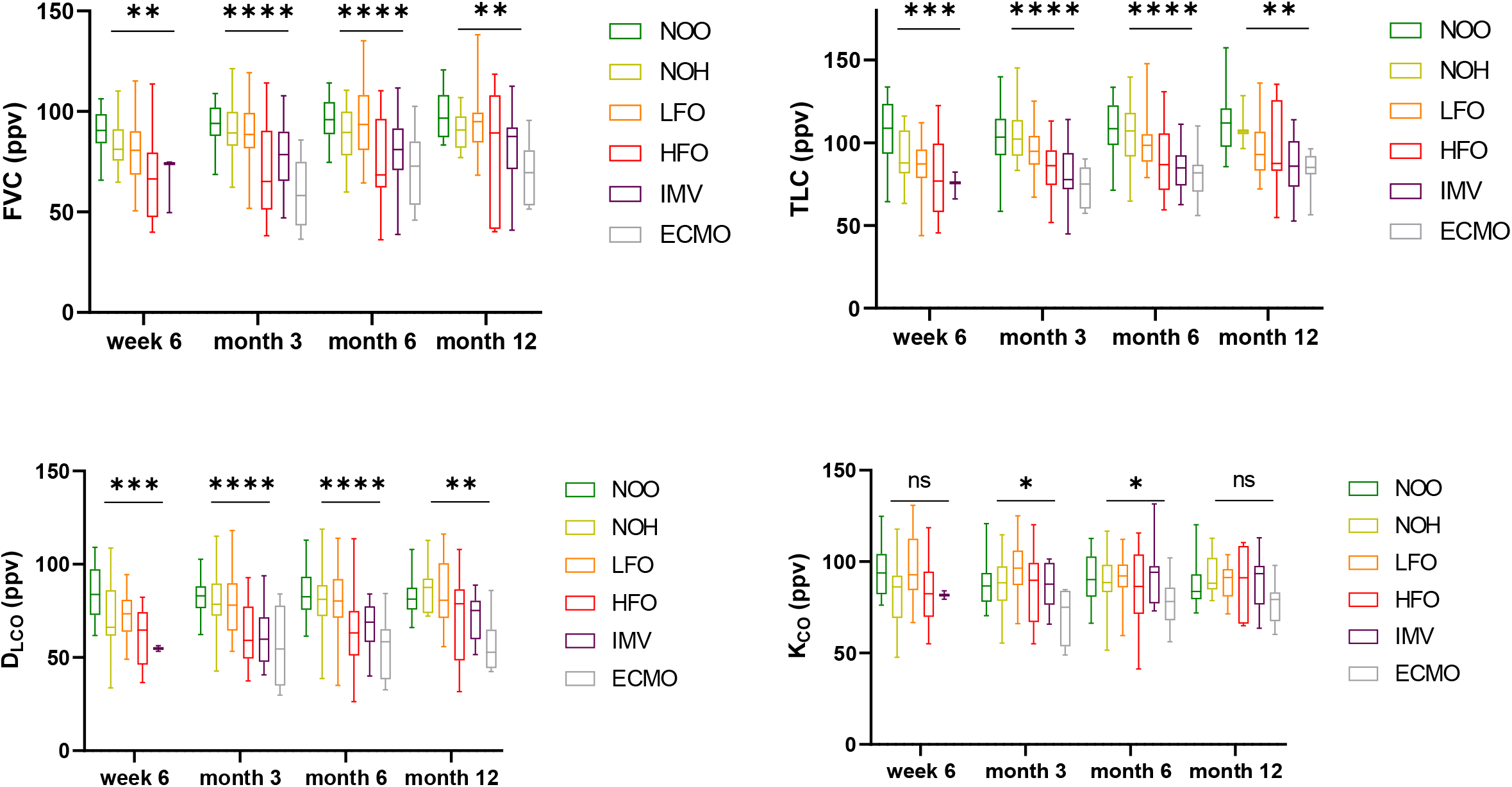
Pulmonary restriction significantly increased with disease severity. Bodyplethysmography 6 weeks, 3, 6 and 12 months post SARS-CoV-2 infection showed significant differences for FVC and TLV for all time points. Diffusion capacity is reduced in the early reconvalescent phase post COVID-19. DLCO significantly correlates with disease severity in the follow-up phase for all time points, whereas KCO is only significantly reduced 3 and 6 months after acute COVID-19. (Abbreviations: ppv – percent predicted value; NOO – no oxygen outpatient; NOH – no oxygen hospitalized, LFO – low-flow oxygen; HFO – high-flow oxygen; IMV – invasive mechanical ventilation; ECMO – extracorporeal membrane oxygenation; ns = P ≥ 0.05; ^*^ = P < 0.05; ^**^ = P < 0.01; ^***^ = P < 0.001; ^****^ < P ≤ 0.0001)

Likewise, impaired D_LCO_ was associated with disease severity. Significant differences were seen in patients stratified by level of respiratory support as a proxy of acute COVID-19 severity (Figure 1). With regard to K_CO_ (D_LCO_ /V_A_, Krogh-Index), a different pattern was observed; significant differences were only seen at either month 3 or 6 post SARS-CoV-2 infection and differences between severity were less pronounced (Figure 1).

There was no association between pulmonary obstruction and disease severity after acute COVID-19 (Table 1). Reduced FEV1 was attributable to concurrent reduced FVC (Figure s1a). Although there were individual cases with reduced airway occlusion pressure (P0.1) and inspiratory muscle strength (Pimax), no statistically significant differences regarding P0.1 and Pimax were seen between severity groups. (Figure s1b).

#### Pulmonary function during follow-up

Patients with pathological pulmonary function in the early post-acute phase (defined as TLC/FVC/D_LCO_ < Lower Limit of Normal (LLN) at first follow-up) showed improvement up to month 12 for pulmonary restriction or reduced D_LCO_ (Figure 2). When all patients were analysed over time, median TLC, FVC, D_LCO_ and K_CO_ increased up to month 6, with no further improvement seen between month 6 and 12 (Figure s2, Table s1).

**Figure 2:**
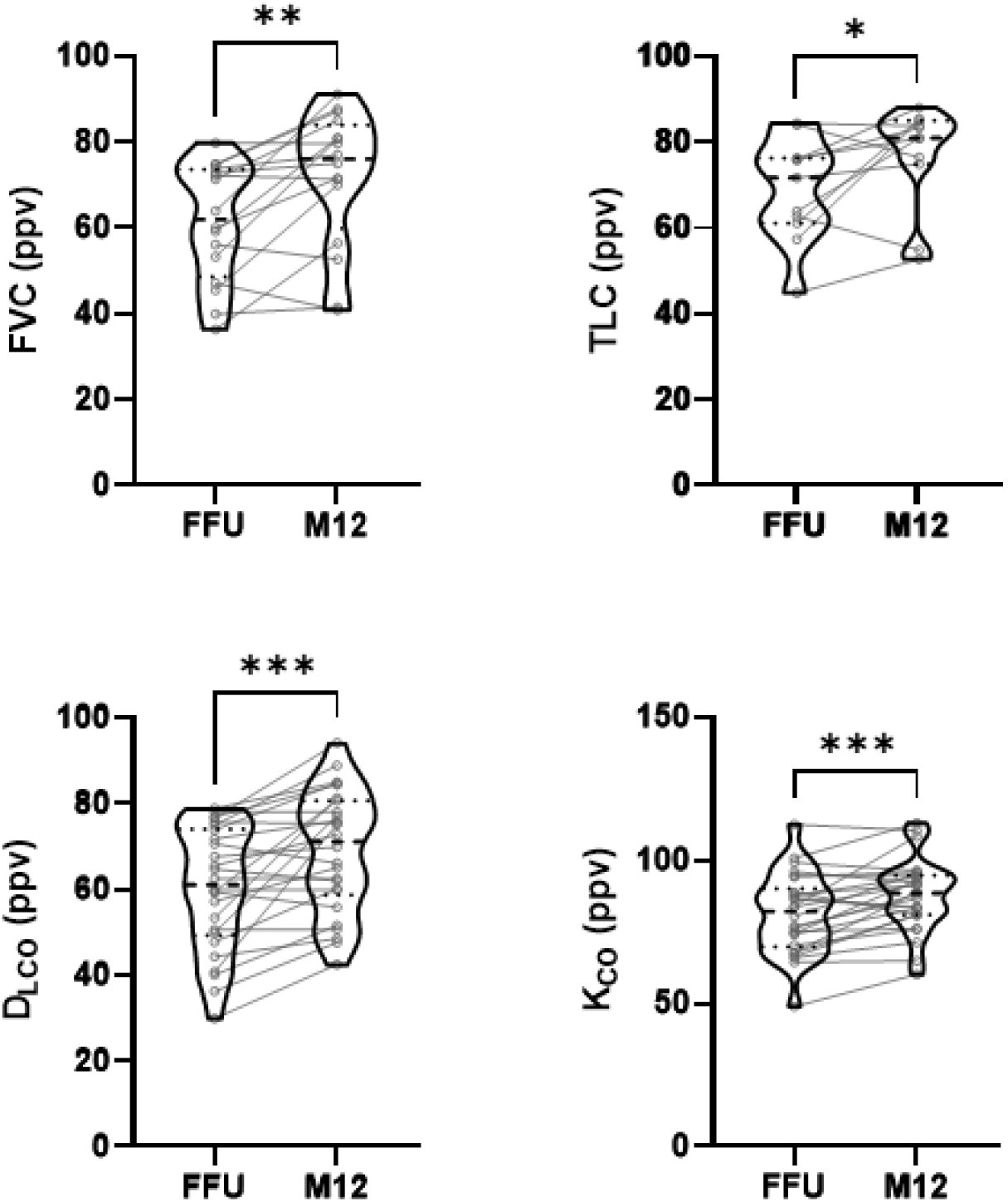
Pulmonary restriction and impaired diffusion capacity improves over time in patients with abnormal pulmonary function test from first follow-up up to month 12. FVC, TLC, DLCO and KCO during follow up showed significant improvements in patients with initially reduced pulmonary function test results. (Abbreviations: FFU – first follow-up, M12 – month 12 follow-up)

### Chest CT

Chest-CT was performed in 88 (51.2%) patients during acute-COVID-19 infection. Median time of first chest CT was 9 days (IQR 6-14) post symptom onset. The CT score assessing lung involvement as suggested by Pan et al. increased significantly with acute COVID-19 severity (p<0.0001) (Figure 3). There was a significant correlation between pulmonary involvement during the acute phase and reduced pulmonary function after acute COVID-19 for TLC, FVC and D_LCO_, (p<0.0001; 0.005; 0.006 respectively) but not for K_CO_ (p=0.244) (Figure 4). Linear regression analysis showed a 15% decrease in TLC and 10% decrease in FVC and D_LCO_ (ppv) for every 10 point increase in CT-chest score during acute SARS-CoV-2 infection.

**Figure 3:**
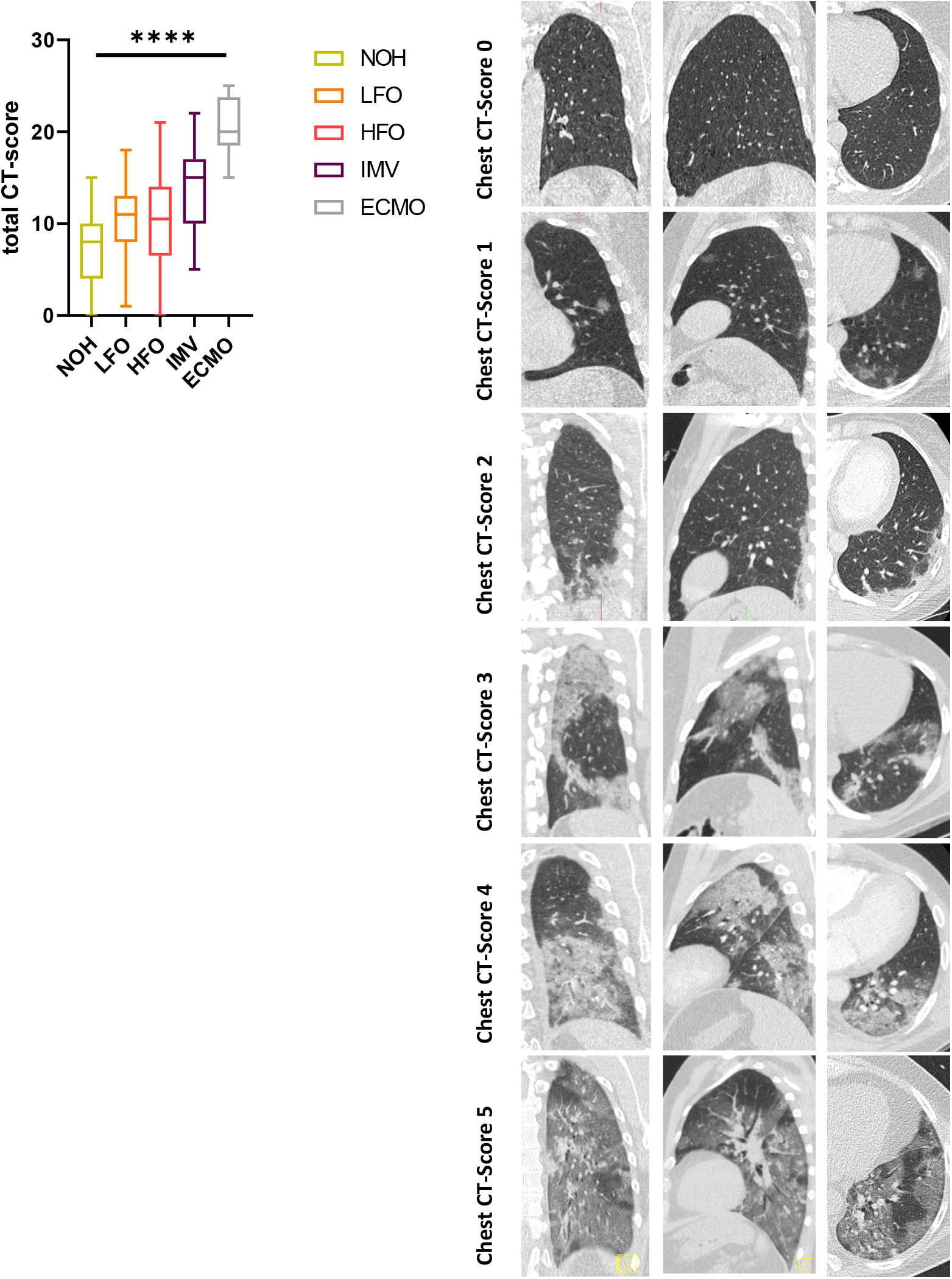
a) CT-score as suggested by Pan et al. at time of hospital admission (median 9 days post symptom onset) showed a significant increase in pulmonary involvement with higher disease severity determined be level of respiratory support. b) Representative CT-chest scans assessed using the 6-point scale of Pan et al. This figure shows left lower lobe involvement of 0% (score 0), <5% (score 1), 5-25% (score 2), 26-50% (score 3), 51-75% (score 4), and >75% (score 5) on axial, coronal, and sagittal CT sections.

**Figure 4:**
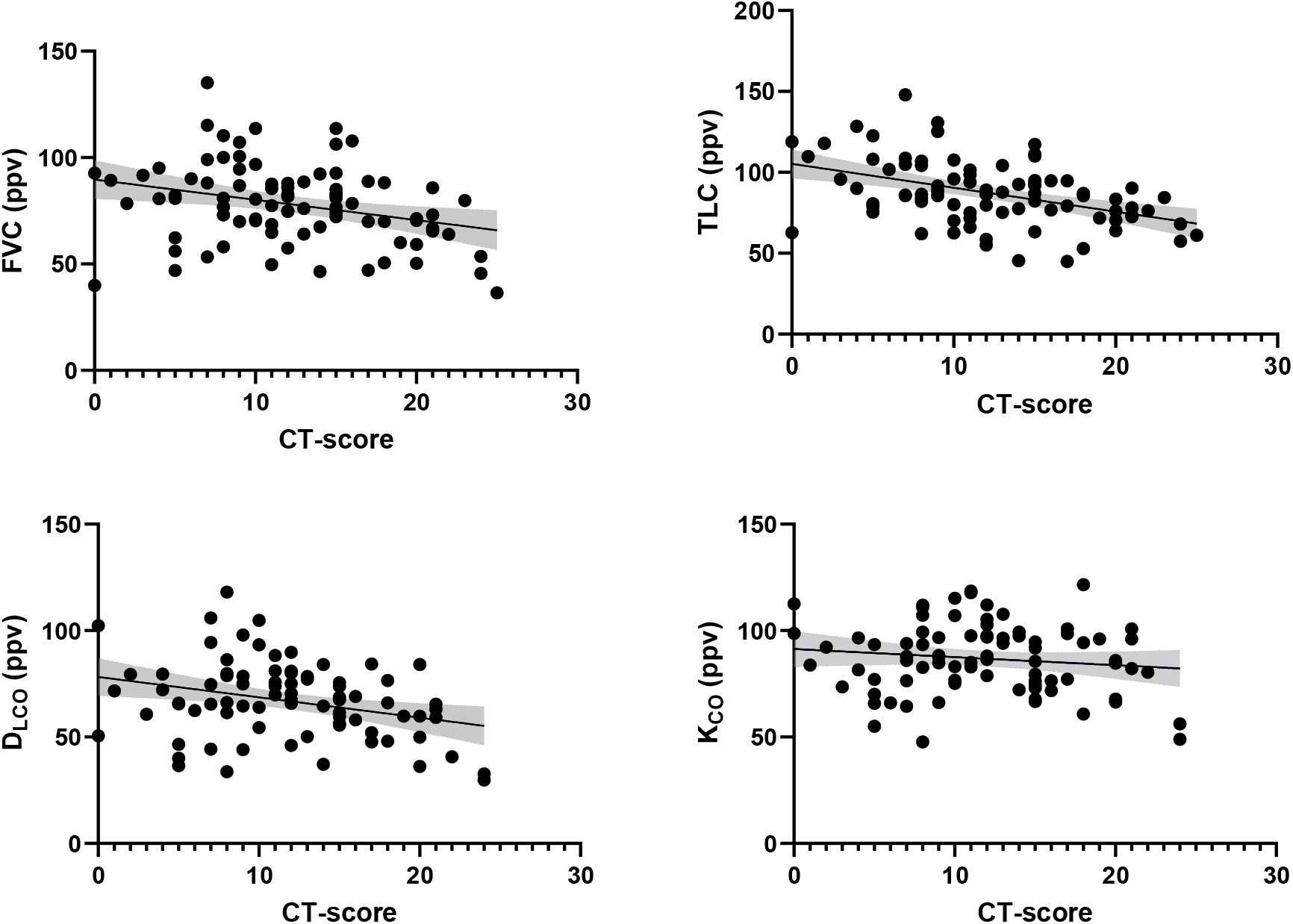
The proportion of pulmonary involvement during acute phase negatively correlates with first pulmonary function test post-acute COVID-19 for TLC, FVC and DLCO. Linear regression analysis reveals for every 10 points increase in CT-score an estimated decrease of 15% TLC and 10% FVC and DLCO post-acute COVID-19.

### Symptom assessment and health related quality of life

The five most common reported symptoms during follow up were fatigue, dyspnea, cough, cognitive impairment and joint pain for all time-points. Symptom load was still high at month 12, with 60.87% of all patients reporting fatigue, 43.48% reporting shortness of breath and 23.19% claiming persistent cognitive impairment (Figure 5a).

**Figure 5:**
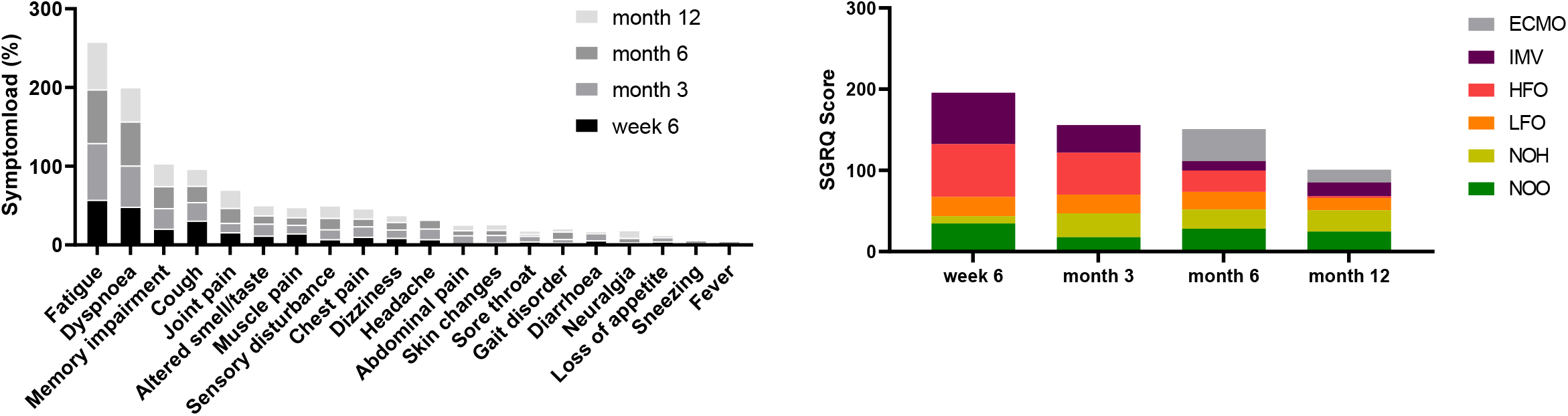
Symptom load and patient reported health outcome: The proportion of patients reporting fatigue, pulmonary- and neurocognitive sequelae remains high 12 months post-acute COVID-19. Median total SGRQ is higher after HFO, IMV and ECMO treatment and decreases until month 12, whereas for LFO, NOH and NOO is remains constant over time.

Standardized assessment of respiratory symptoms by SGRQ showed an improvement over 12 months post symptom onset in patients with higher disease severity in the acute phase (ECMO, IMV, HFO), whereas SGRQ scores in patients in lower severity categories (LFO, NOH and NOO) remained almost constant during 12 months of follow-up. (Figure 5b).

In general, significant correlations with total SGRQ score were observed for FVC (p<0.0001), D_LCO_ (p<0.001) and K_CO_ (p<0.0001) but not for TLC (p=0.091) (Table 2, Figure s5). The contribution of SGRQ sub-scores for symptoms, activity and impact are shown in Table 2.

**Table 2:** Patient reported outcome post-acute COVID-19 correlates with pulmonary function outcome. SGRQ results of all time-points were matched with respective pulmonary function tests. Total and SGRQ sub-scores negatively correlate with D_LCO_, K_CO_ and FVC, however not significantly with TLC.

| SGRQ |  |  |  |  |  |  |  |  |  |  |  |  |
| --- | --- | --- | --- | --- | --- | --- | --- | --- | --- | --- | --- | --- |
|  | *Total* |  |  | *Symptoms* |  |  | *Activity* |  |  | *Impact* |  |  |
| PFT | *r* | *R*<sup>2</sup> | *p* | *r* | *R*<sup>2</sup> | *p* | *r* | *R*<sup>2</sup> | *p* | *r* | *R*<sup>2</sup> | *p* |
| D<sub>LCO</sub> | -0.48 | 0.22 | <0,0001 | -0.37 | 0.14 | <0.0001 | -0.49 | 0.24 | <0.0001 | -0.38 | 0.14 | <0.0001 |
| K<sub>CO</sub> | -0.35 | 0.12 | <0,0001 | -0.29 | 0.08 | 0.0002 | -0.34 | 0.11 | <0.0001 | -0.35 | 0.12 | <0.0001 |
| FVC | -0.41 | 0.17 | <0,0001 | -0.25 | 0.06 | 0.0011 | -0.37 | 0.13 | <0.0001 | -0.35 | 0.12 | <0.0001 |
| TLC | -0.14 | 0.02 | 0.0911 | -0.07 | 0.01 | 0.3694 | -0.15 | 0.02 | 0.0519 | -0.10 | 0.01 | 0.2130 |

### Risk factors for pulmonary restriction and reduced diffusion capacity

Univariate logistic regression showed an association of pulmonary restriction and reduced D_LCO_ with disease severity, gender, SGRQ outcome (score>25), Charlson Comorbidity Index and cardiovascular disease (Table 3). The odds of restriction were 1.7 (95% CI 1.37-2.15, p=0.0001) times higher for every single increment in disease severity category (i.e. from LFO to HFO) and 1.8 (1.19-2.74, p=0.01) for every 5 points increase in CT-score. For reduced D_LCO_, logistic regression showed no significant association with initial pulmonary involvement (p=0.11), but for disease severity (OR 1.49, 95% CI 1.20-1.84; p<0.0001). Effect sizes were also adjusted for age, sex, BMI and disease severity (Table 3).

**Table 3:** Association of demographic characteristics, clinical indicators and comorbidities with pulmonary restriction and impaired D_LCO_ post-acute COVID-19. Univariate analysis revealed male gender, disease severity, SGRQ score >25, Charlson Comorbidity Index and cardiovascular disease to be associated with pulmonary restriction and reduced D_LCO_. A relationship between CT-chest score was only seen for patients developing restriction. In multivariable analysis, adjustment for age, BMI, gender and disease severity showed SGRQ outcome over the threshold of 25 to be associated with both pulmonary restriction and impaired D_LCO_. Patient characteristics and comorbidities were collected at study inclusion. Worst SGRQ outcome independent of follow-up was used for univariate and multivariate analysis.

|  | Pulmonary restriction |  |  |  | Impaired D<sub>LCO</sub> |  |  |  |
| --- | --- | --- | --- | --- | --- | --- | --- | --- |
|  | OR (95% CI) | p | aOR (95% CI) | p | OR (95% CI) | p | aOR (95% CI) | p |
| Age (per 10yrs) | 1.21 (0.96-1.51) | 0.101 |  |  | 1.14 (0.92-1.41) | 0.242 |  |  |
| BMI>30 | 1.55 (0.79-3.04) | 0.201 |  |  | 0.73 (0.38-1.49) | 0.337 |  |  |
| Gender (male) | 4.52 (2.11-9.68) | < 0.0001 |  |  | 2.14 (1.15-3.96) | 0.016 |  |  |
| Disease severity | 1.72 (1.37-2.15) | < 0.0001 |  |  | 1.49 (1.20-1.84) | < 0.0001 |  |  |
| CT-chest (scale) | 1.80 (1.19-2.74) | 0.006 | 1.42 (0.85-2.36) | 0.176 | 1.42 (0.92-2.19) | 0.111 | 0.91 (0.50-1.64) | 0.743 |
| SGRQ>25 | 2.63 (1.22-5.67) | 0.021 | 2.80 (1.14-6.91) | 0.025 | 4.91 (2.28-10.58) | < 0.0001 | 5.99 (2.47-14.53) | < 0.0001 |
| CCI | 1.48 (1.06-2.07) | 0.023 | 1.25 (0.71-2.18) | 0.437 | 1.54 (1.10-2.15) | 0.013 | 2.07 (1.11-3.85) | 0.021 |
| Cardiovascular dis. | 2.06 (1.09-3.88) | 0.026 | 1.21 (0.55-2.67) | 0.644 | 1.51 (0.82-2.79) | 0.190 | 1.14 (0.53-2.46) | 0.736 |
| Pulmonary dis. | 1.40 (0.58-3.43) | 0.447 | 1.25 (0.45-3.46) | 0.663 | 0.70 (0.20-1.62) | 0.700 | 0.62 (0.24-1.58) | 0.312 |
| Renal dis. | 1.75 (0.68-4.50) | 0.244 | 1.38 (0.47-4.02) | 0.561 | 2.03 (0.71-5.90) | 0.187 | 1.85 (0.59-5.77) | 0.293 |
| Diabetes | 1.80 (0.85-3.78) | 0.121 | 0.90 (0.39-2.07) | 0.798 | 1.78 (0.78-4.07) | 0.172 | 1.29 (0.53-3.17) | 0.578 |
| Liver dis. | 0.83 (0.42-1.64) | 0.584 | 0.74 (0.68-1.95) | 0.536 | 1.17 (0.68-2.01) | 0.578 | 1.01 (0.80-1.28) | 0.907 |
| Organ transplant | 0.06 (0.07-6.49) | 0.722 | 0.67 (0.06-7.68) | 0.745 | 1.93 (0.20-18.88) | 0.574 | 1.90 (0.19-19.27) | 0.588 |
| Autoimmune dis. | 0.89 (0.53-1.48) | 0.639 | 0.86 (0.44-1.71) | 0.676 | 1.40 (0.55-3.57) | 0.485 | 1.29 (0.57-2.80) | 0.574 |
| Immunosuppression | 3.20 (0.86-11.81) | 0.081 | 2.59 (0.64-10.48) | 0.182 | 1.49 (0.37-5.96) | 0.575 | 1.05 (0.25-4.40) | 0.945 |
| Smoking | 1.63 (0.85-3.15) | 0.142 | 1.31 (0.62-2.77) | 0.480 | 0.99 (0.52-1.89) | 0.981 | 0.74 (0.36-1.53) | 0.981 |

## Discussion

In this study of COVID-19 survivors, we longitudinally analysed pulmonary function, respiratory symptoms and health related quality of life and studied CT chest morphology at acute phase of 180 patients during 12 months after SARS-CoV-2 infection. We identified demographic characteristics, clinical indicators and comorbidities that increase the risk and severity of pulmonary injury. The detailed data on pulmonary function presented gives first insight into different patterns of pulmonary impairment according to clinical severity in the acute phase and its sequelae up to 12 months post SARS-CoV-2 infection.

Reduced FVC, TLC and D_LCO_ were associated with severe and critical COVID-19 in the literature, representing patients with LFO, HFO, IMV and ECMO in our cohort ^12, 15, 39^. This study demonstrates that the degree of pulmonary functional impairment correlates with clinical severity during acute COVID-19 and that these differences in pulmonary function were still apparent after 12 months of follow-up.

Pulmonary restriction was associated with the degree of lung parenchymal involvement seen on CT scans during acute COVID-19, reflecting inflammation and fibrotic transformation following SARS-CoV-2 infection. Increasing evidence suggests a profibrotic phenotype following SARS-CoV-2 infection ^11, 1, 6, 26^, in line with other viral causes of pneumonia such as SARS, MERS and influenza infections ^17, 14, 28^. Post mortem analysis of lung tissue in lethal COVID-19 were reported to show ultrastructural alteration including alveolar collapse and fibrosis ^1, 26^. Also, similarities in gene expression between idiopathic pulmonary fibrosis and COVID-19 in explanted lungs of patients undergoing lung transplantations or post mortem analysis were found using single-cell RNA sequencing, including Keratin-17 expressing epithelial cells, profibrotic macrophages and myofibroblasts ^18, 1^. Thus, analysis of CT scans during the acute phase may have prognostic relevance for patients.

It could be argued that pulmonary restriction after acute COVID-19 is caused by ventilator-induced lung injury (VILI), a common observation in ARDS patients ^31, 34^ including subsequent pulmonary restriction and reduced D_LCO_ ^24, 35^. In this study however, there was no obvious difference in FVC, TLC and D_LCO_ at all follow-up visits between patients who needed mechanical ventilation and those who received high-flow oxygen therapy. Although more data is needed to confirm this hypothesis, our data indicate that post COVID-19 pulmonary restriction is probably not caused by VILI, but rather by consequences of viral infection.

Two different types of restriction were discernible in this study population: the pattern of simple pulmonary restriction was more frequently observed in patients with higher initial disease severity, whereas complex pulmonary restriction was seen predominantly in patients with less severe COVID-19. Complex restriction, according to the definition by Clay et al. ^4^, describes a restriction pattern where FVC is disproportionally reduced compared to the reduction of TLC, combined with an increased RV and RV/TLC ratio, and usually without evidence of obstruction. Complex restriction occurs in obesity or may indicate occult obstruction, but also typically can be observed in neuromuscular diseases. Whether particularly the latter condition is related to the complex restriction pattern after acute COVID-19 in this study population warrants further investigation. The simple restriction pattern observed in severely ill patients may reflect fibrotic changes in the lung interstitium typically associated with ARDS.

In line with previous observations of impaired carbon monoxide diffusion capacity post-acute COVID-19 at time of hospital discharge ^2, 22, 25^, D_LCO_, but not K_CO_ (D_LCO_ / V_A_) was reduced and significantly different among patients of all levels of acute COVID-19 severity. However, patients requiring ECMO treatment in the acute phase of COVID-19 still had reduced D_LCO_ as well as K_CO_ 3, 6 and 12 months post infection. Together with the distribution of simple vs. complex restriction patterns described above, we hypothesize that in milder disease a loss of ventilated alveolar units is the dominant phenomenon of respiratory impairment, whereas in the most severe cases diffusion is impaired by pathologic processes in the alveolo-capillary barrier such as interstitial fibrosis.

Analysis of pulmonary function over time revealed three groups: (i) those less severely affected during acute COVID-19 who did not show significant alterations in pulmonary function during follow-up, (ii) those with compromised pulmonary function at first follow-up who showed improvement over time and (iii) those with severely compromised pulmonary function at first follow-up who did not show relevant improvement over time. In particular, many patients with the most severe respiratory failure who needed ECMO treatment still have significant pulmonary impairment one year after the acute disease.

The symptom cluster including fatigue, dyspnoea and cognitive deficits as described for the early convalescent phase ^23^ persisted over 12 months of follow-up in our study population. Respiratory health related quality of life as captured by total SGRQ improved over time, but with a relevant proportion of patients remaining above the threshold value of 25 one year after acute COVID-19. TLC however did not correlate with SGRQ score, likely due to the high proportion of patients with complex restriction and preserved TLC.

Limitations of this study were the availability of data from a single centre at this point of time, and the reduced number of patients available in the first and the last 12 month follow-up visit, particularly in the group of patients after invasive mechanical ventilation and ECMO treatment.

By summarizing the results from pulmonary function tests with assessment of respiratory symptoms and the evolution of findings over time, we hypothesize that two main patterns of pulmonary involvement are discernible after COVID-19: in patients with severe disease and particularly those with respiratory failure requiring ECMO treatment, a pattern of interstitial lung involvement characterized by simple restriction and reduction of diffusion capacity predominates. This pattern has potential for functional and subjective improvement over time during the first year of follow-up. In patients with mild to moderate initial disease however, a disease pattern characterized by a loss of ventilated area and symptom persistence over one year after follow-up predominates. Particularly for the latter pattern, potential underlying mechanisms are unknown, and these patterns of pulmonary injury will need to be confirmed and further characterized in larger and multi-centric studies.

In conclusion, this study demonstrated the relevance of initial disease severity and results of thoracic CT for pulmonary functional impairment and respiratory symptoms in the first year after SARS-CoV-2 infection in hospitalized patients.

## Data Availability

The data that support the findings of this study are available from the corresponding author upon reasonable request.

## Author contributions

F.S. and T.Z. had full access to all of the data in the study and take responsibility for the integrity of the data and the accuracy of the data analysis. F.S., T.Z. and M.W. developed the study design, performed data analysis and interpretation and wrote the manuscript. F.D. and T.P. performed data analysis and interpretation of radiological data. C.T., C.R.G., F.A., P.K., R.M.R., M.M., L.J.L, E.T.H., F.G., B.T.W., N.S., F.K., L.E.S. contributed substantially to the study design data analysis and interpretation, and the writing of the manuscript.

## Acknowledgement

We thank the Pa-COVID-19 study group with Stefan Hippenstiel, Pinkus Tober-Lau, Sascha S. Haenel, David Hillus, Tilman Lingscheid, Holger Müller-Redetzky, Alexander Uhrig, Miriam S. Stegemann, Ralf H. Hübner, Kai-Uwe Eckardt, Martin Möckel, Felix Balzer, Claudia Spies, Steffen Weber-Karstens, Frank Tacke, Chantip Dang-Heine, Michael Hummel, Georg Schwanitz, Uwe D. Behrens, Maria Rönnefarth, Sein Schmidt, Alexander Krannich, Christof von Kalle, Victor M. Corman and Christian Drosten that contributed to set up and realization of the study platform; Linda Jürgens, Malte Kleinschmidt, Sophy Denker, Moritz Pfeiffer, Belén Millet-Pascual-Leone, Felix Machleidt, Sebastian Albus, Felix Bremer, Jan-Moritz Doehn, Tim Andermann, Carmen Garcia, Philipp M. Krause, Liron Lechtenberg, Yaosi Li, Panagiotis Pergantis, Till Jacobi, Teresa Ritter, Berna Yedikat, Lennart Pfannkuch, Christian Zobel, Ute Kellermann, Susanne Fieberg, Laure Bosquillon de Jarcy, Anne Wetzel, Christoph Tabeling, Markus Brack, Jan M. Kruse, Daniel Zickler, Andreas Edel, Britta Stier, Roland Körner, Nils B. Mueller and Philipp Enghard to obtaining informed consent and biosamples and Nadine Olk, Willi M. Koch, Saskia Zvorc, Lucie Kretzler, Lil A. Meyer-Arndt, Linna Li and Isabelle Wirsching, Paula Stubbemann, Jo Bagli, Olivia Zielonka and Vivien Schreiber to data collection.

M.M. is participant in the BIH-Charité Digital Clinician Scientist Program funded by the Charité –Universitätsmedizin Berlin, the Berlin Institute of Health, and the German Research Foundation (DFG). M.W. is supported by grants from the German Research Foundation, SFB-TR84 C6 and C9, SFB 1449 B2, by the German Ministry of Education and Research (BMBF) in the framework of CAPSyS (01ZX1304B), CAPSyS-COVID (01ZX1604B), SYMPATH (01ZX1906A), PROVID (01KI20160A), P4C (16GW0141), MAPVAP (16GW0247), NUM-NAPKON (01KX2021), and by the Berlin Institute of Health (CM-COVID).

## Literature

1. Bharat A, Querrey M, Markov NS, et al. Lung transplantation for patients with severe COVID-19. Science Translational Medicine (2020); 12(574): eabe4282.

2. Chapman DG, Badal T, King GG and Thamrin C Caution in Interpretation of Abnormal Carbon Monoxide Diffusion Capacity in COVID-19 Patients. European Respiratory Journal (2020): 2003263.

3. Chen R, Gao Y, Chen M, et al. Impaired pulmonary function in discharged patients with COVID-19: more work ahead. European Respiratory Journal (2020); 56(1): 2002194.

4. Clay RD, Iyer VN, Reddy DR, Siontis B and Scanlon PD The “Complex Restrictive” Pulmonary Function Pattern: Clinical and Radiologic Analysis of a Common but Previously Undescribed Restrictive Pattern. Chest (2017); 152(6): 1258–1265.

5. Criée C-P, Baur X, Berdel D, et al. Leitlinie zur Spirometrie. Pneumologie (2015); 69(03): 147–164.

6. Ferreira-Gomes M, Kruglov A, Durek P, et al. In severe COVID-19, SARS-CoV-2 induces a chronic, TGF-β-dominated adaptive immune response. medRxiv (2020): 2020.2009.2004.20188169.

7. Gelpi M, Argentiero J, Jones PW and Ronit A A Scoring Application for the St. George’s Respiratory Questionnaire. Chest (2016); 150(3): 747–748.

8. Graham BL, Brusasco V, Burgos F, et al. 2017 ERS/ATS standards for single-breath carbon monoxide uptake in the lung. European Respiratory Journal (2017); 49(1): 1600016.

9. Graham BL, Brusasco V, Burgos F, et al. DLCO: adjust for lung volume, standardised reporting and interpretation. European Respiratory Journal (2017); 50(2): 1701144.

10. Graham BL, Steenbruggen I, Miller MR, et al. Standardization of Spirometry 2019 Update. An Official American Thoracic Society and European Respiratory Society Technical Statement. American Journal of Respiratory and Critical Care Medicine (2019); 200(8): e70–e88.

11. Gralinski LE, Bankhead A, 3rd, Jeng S, et al. Mechanisms of severe acute respiratory syndrome coronavirus-induced acute lung injury. mBio (2013); 4(4).

12. Guler SA, Ebner L, Aubry-Beigelman C, et al. Pulmonary function and radiological features 4 months after COVID-19: first results from the national prospective observational Swiss COVID-19 lung study. European Respiratory Journal (2021); 57(4): 2003690.

13. Guler SA, Ebner L, Beigelman C, et al. Pulmonary function and radiological features four months after COVID-19: first results from the national prospective observational Swiss COVID-19 lung study. European Respiratory Journal (2021): 2003690.

14. Hsieh M-J, Lee W-C, Cho H-Y, et al. Recovery of pulmonary functions, exercise capacity, and quality of life after pulmonary rehabilitation in survivors of ARDS due to severe influenza A (H1N1) pneumonitis. Influenza Other Respir Viruses (2018); 12(5): 643–648.

15. Huang C, Huang L, Wang Y, et al. 6-month consequences of COVID-19 in patients discharged from hospital: a cohort study. The Lancet (2021); 397(10270): 220–232.

16. Huang Y, Tan C, Wu J, et al. Impact of coronavirus disease 2019 on pulmonary function in early convalescence phase. Respiratory Research (2020); 21(1): 163.

17. Hui DS, Joynt GM, Wong KT, et al. Impact of severe acute respiratory syndrome (SARS) on pulmonary function, functional capacity and quality of life in a cohort of survivors. Thorax (2005); 60(5): 401–409.

18. Iyonaga K, Miyajima M, Suga M, Saita N and Ando M Alterations in cytokeratin expression by the alveolar lining epithelial cells in lung tissues from patients with idiopathic pulmonary fibrosis. J Pathol (1997); 182(2): 217–224.

19. Kurth F, Roennefarth M, Thibeault C, et al. Studying the pathophysiology of coronavirus disease 2019: a protocol for the Berlin prospective COVID-19 patient cohort (Pa-COVID-19). Infection (2020); 48(4): 619–626.

20. Laszlo G Standardisation of lung function testing: helpful guidance from the ATS/ERS Task Force. Thorax (2006); 61(9): 744–746.

21. Lerum TV, Aaløkken TM, Brønstad E, et al. Dyspnoea, lung function and CT findings 3 months after hospital admission for COVID-19. European Respiratory Journal (2021); 57(4): 2003448.

22. Mo X, Jian W, Su Z, et al. Abnormal pulmonary function in COVID-19 patients at time of hospital discharge. European Respiratory Journal (2020); 55(6): 2001217.

23. Nalbandian A, Sehgal K, Gupta A, et al. Post-acute COVID-19 syndrome. Nat Med (2021); 27(4): 601–615.

24. Neff TA, Stocker R, Frey H-R, Stein S and Russi EW Long-term Assessment of Lung Function in Survivors of Severe ARDSa. Chest (2003); 123(3): 845–853.

25. Nusair S Abnormal carbon monoxide diffusion capacity in COVID-19 patients at time of hospital discharge. European Respiratory Journal (2020); 56(1): 2001832.

26. Ochs M, Timm S, Elezkurtaj S, et al. Collapse induration of alveoli is an ultrastructural finding in a COVID-19 patient. European Respiratory Journal (2021): 2004165.

27. Pan F, Ye T, Sun P, et al. Time Course of Lung Changes at Chest CT during Recovery from Coronavirus Disease 2019 (COVID-19). Radiology (2020); 295(3): 715–721.

28. Park WB, Jun KI, Kim G, et al. Correlation between Pneumonia Severity and Pulmonary Complications in Middle East Respiratory Syndrome. J Korean Med Sci (2018); 33(24): e169.

29. Pellegrino R, Viegi G, Brusasco V, et al. Interpretative strategies for lung function tests. Eur Respir J (2005); 26(5): 948–968.

30. Polak SB, Van Gool IC, Cohen D, Von Der Thüsen JH and Van Paassen J A systematic review of pathological findings in COVID-19: a pathophysiological timeline and possible mechanisms of disease progression. Modern Pathology (2020); 33(11): 2128–2138.

31. Ricard JD, Dreyfuss D and Saumon G Ventilator-induced lung injury. European Respiratory Journal (2003); 22(42 suppl): 2s–9s.

32. Sonnweber T, Sahanic S, Pizzini A, et al. Cardiopulmonary recovery after COVID-19: an observational prospective multicentre trial. European Respiratory Journal (2021); 57(4): 2003481.

33. Stanojevic S, Quanjer P, Miller MR and Stocks J The Global Lung Function Initiative: dispelling some myths of lung function test interpretation. Breathe (2013); 9(6): 462–474.

34. Thompson BT, Chambers RC and Liu KD Acute Respiratory Distress Syndrome. New England Journal of Medicine (2017); 377(6): 562–572.

35. Von Bahr V, Kalzén H, Frenckner B, et al. Long-term pulmonary function and quality of life in adults after extracorporeal membrane oxygenation for respiratory failure. Perfusion (2019); 34(1_suppl): 49–57.

36. Who WHO R&D Blueprint novel Coronavirus COVID-19 Therapeutic Trial Synopsis. R&D Blueprint (2020).

37. Wiersinga WJ, Rhodes A, Cheng AC, Peacock SJ and Prescott HC Pathophysiology, Transmission, Diagnosis, and Treatment of Coronavirus Disease 2019 (COVID-19): A Review. JAMA (2020); 324(8): 782–793.

38. Wölfel R, Corman VM, Guggemos W, et al. Virological assessment of hospitalized patients with COVID-2019. Nature (2020); 581(7809): 465–469.

39. Wu X, Liu X, Zhou Y, et al. 3-month, 6-month, 9-month, and 12-month respiratory outcomes in patients following COVID-19-related hospitalisation: a prospective study. Lancet Respir Med (2021).

40. Zhao Y-M, Shang Y-M, Song W-B, et al. Follow-up study of the pulmonary function and related physiological characteristics of COVID-19 survivors three months after recovery. EClinicalMedicine (2020); 25: 100463.

