## Supplementary material for "Severity of respiratory failure and computed chest tomography in acute COVID-19 correlates with pulmonary function and respiratory symptoms after infection with SARS-CoV-2: an observational longitudinal study over 12 months"

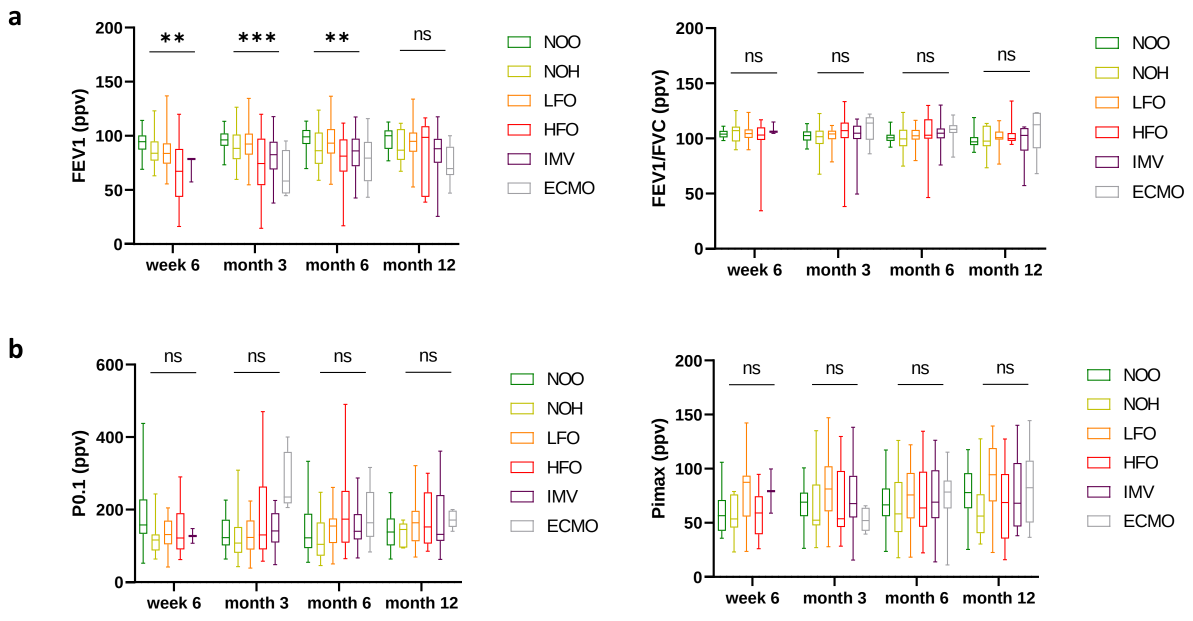


**Figure s1:** Pulmonary obstruction and respiratory muscle testing is independent of COVID disease severity. a) Median Tiffeneau index (FEV1/FVC) remains constant post-acute COVID-19 after different courses of acute infection. FEV1 decease amongst patients with HFO, IMV and ECMO is explained by reduction in FVC (s. Figure 1). b) No statistical significant difference regarding the mechanical index of respiratory drive or airway occlusion pressure (P0.1) and inspiratory muscle strength (Pimax) between different disease groups were detected.

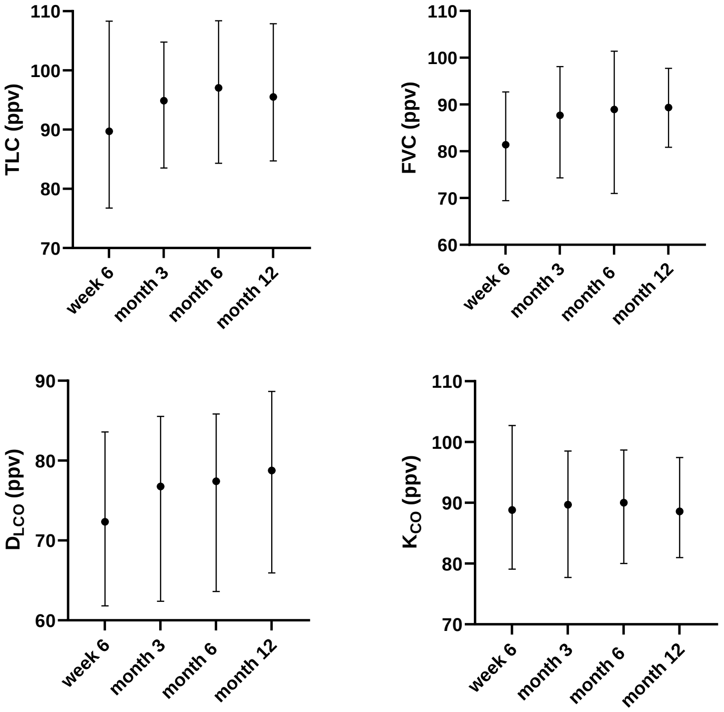


**Figure s2**: Median TLC, FVC and $D_{LCO}$ of all patients improved during early reconvalescence up to month 6, with no further improvement between month 6 and 12. This was not observed for $K_{CO}$.

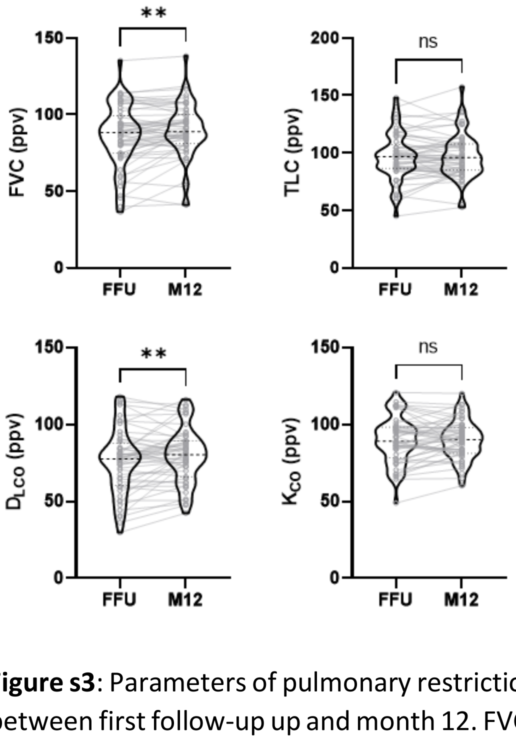


**Figure s3**: Parameters of pulmonary restriction and impaired diffusion capacity over time in all patients between first follow-up up and month 12. FVC and $D_{LCO}$ showed significant improvements in all patients during follow-up until month 12. (Abbreviations: FFU – first follow-up, M12 – month 12 follow-up)

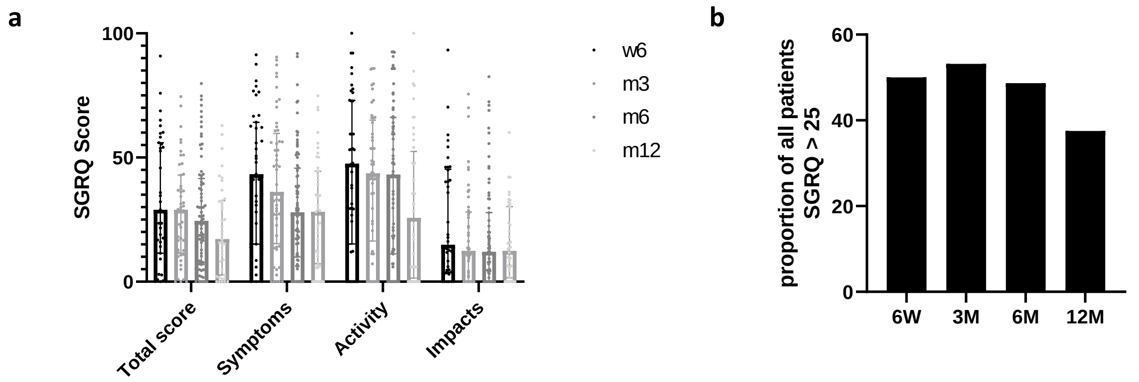


**Figure s4**: SGRQ total and subscore over time: **a)** Within early follow up, no improvement was seen between week 6 and month 3, with a median score of 28.93 [IQR 11.43-55.92] at week 6 post symptom onset and 28.93 [11.51-42.95] at month 3, with a reduction hereafter at month 6 and month 12 (24.48 [6.91-41.58]; 17.14 [2.70-32.59]). **b)** Proportion of all patients with an SGRQ score > 25 over time confirm this trend with a decrease after 6 month follow-up.

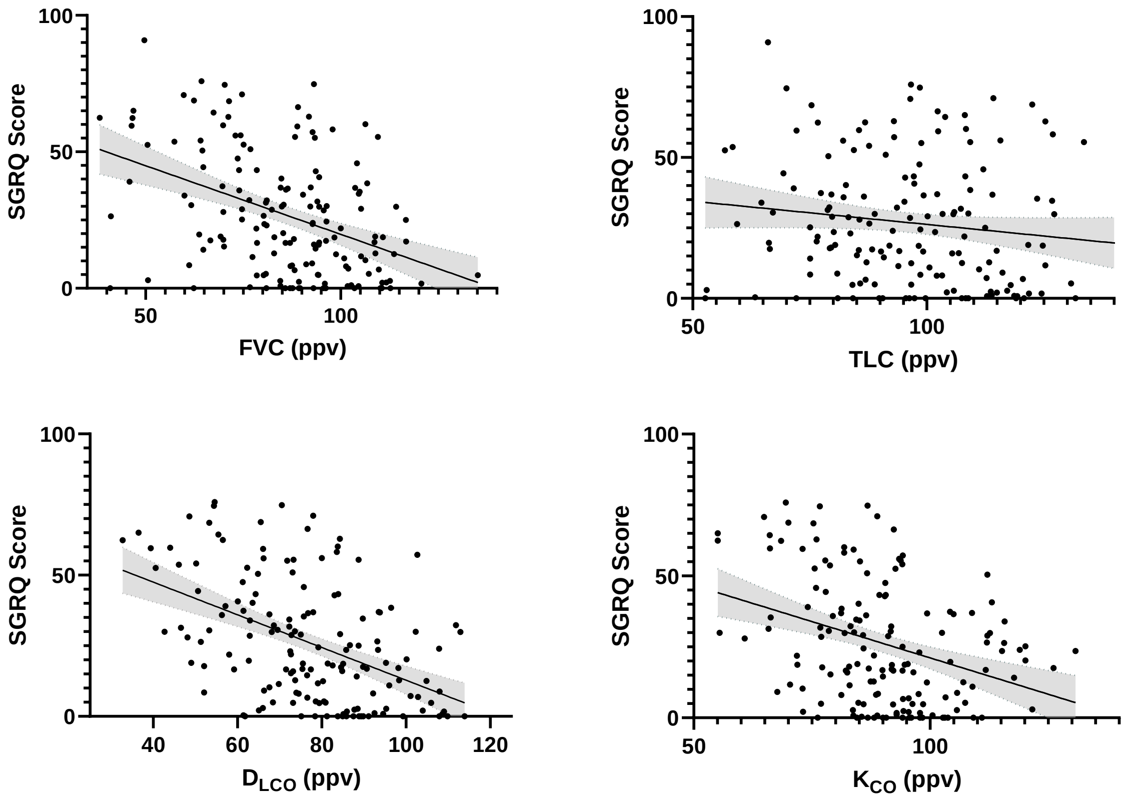


**Figure s5**: Linear regression analysis of total SGRQ Score and pulmonary function post-acute COVID-19: A negative correlation, that is significant for FVC, DLCO and KCO describes a higher SGRQ score in patients with distorted diffusion capacity or reduced FVC. For every 10% improvement in DLCO, KCO and FVC, estimated SGRQ improves by 6, 5 and 5 points respectively. SGRQ results of all time-points were matched with respective pulmonary function tests.

| | NOO | | | NOH | | | LFO | | | HFO | | | IMV | | | ECMO | | |
|---|---|---|---|---|---|---|---|---|---|---|---|---|---|---|---|---|---|---|
| DLCO w6 | ***83.79*** | 97.27 | 72.81 | ***66.17*** | 86.11 | 61.63 | ***73.49*** | 80.99 | 63.81 | ***65.55*** | 74.76 | 48.30 | ***53.26*** | 56.29 | 46.27 | | | |
| DLCO m3 | ***83.00*** | 88.24 | 76.49 | ***78.53*** | 89.73 | 72.41 | ***78.27*** | 90.15 | 65.35 | ***61.27*** | 78.59 | 48.59 | ***58.88*** | 71.18 | 49.37 | ***54.57*** | 77.86 | 34.89 |
| DLCO m6 | ***82.66*** | 93.53 | 77.77 | ***81.23*** | 87.73 | 71.27 | ***80.31*** | 92.89 | 70.77 | ***62.67*** | 78.43 | 52.30 | ***67.56*** | 77.42 | 58.37 | ***59.04*** | 63.61 | 36.19 |
| DLCO m12 | ***82.77*** | 90.36 | 77.245 | ***88.425*** | 93.67 | 74.52 | ***78.09*** | 103.285 | 66.80 | ***78.75*** | 96.26 | 49.53 | ***75.21*** | 79.09 | 59.98 | ***58.96*** | 75.46 | 44.98 |
| KCO w6 | ***93.78*** | 104.19 | 82.14 | ***86.16*** | 92.32 | 69.22 | ***92.86*** | 112.61 | 84.43 | ***78.81*** | 95.55 | 69.73 | ***84.03*** | 93.06 | 79.43 | | | |
| KCO m3 | ***86.685*** | 93.782 | 78.00 | ***88.38*** | 97.46 | 78.547 | ***96.58*** | 105.75 | 87.55 | ***88.98*** | 100.00 | 65.00 | ***89.70*** | 98.55 | 76.52 | ***75.01*** | 84.03 | 53.70 |
| KCO m6 | ***90.20*** | 103.25 | 80.52 | ***89.61*** | 98.67 | 83.24 | ***92.12*** | 99.05 | 85.90 | ***88.17*** | 107.45 | 73.36 | ***94.64*** | 98.54 | 77.17 | ***74.14*** | 84.89 | 66.42 |
| KCO m12 | ***84.14*** | 97.06 | 78.565 | ***89.095*** | 103.00 | 88.11 | ***87.40*** | 95.33 | 79.88 | ***97.04*** | 109.51 | 72.85 | ***94.37*** | 100.89 | 83.28 | ***81.20*** | 90.56 | 66.5 |
| FEV1 w6 | ***95.54*** | 100.73 | 89.26 | ***83.99*** | 94.97 | 77.02 | ***83.69*** | 93.08 | 74.20 | ***73.08*** | 89.87 | 43.31 | ***68.04*** | 78.63 | 55.68 | | | |
| FEV1 m3 | ***96.22*** | 102.17 | 90.83 | ***88.43*** | 101.28 | 78.44 | ***91.92*** | 101.79 | 81.04 | ***76.92*** | 101.53 | 53.17 | ***77.46*** | 90.67 | 65.25 | ***59.58*** | 89.64 | 46.08 |
| FEV1 m6 | ***99.69*** | 105.44 | 92.02 | ***85.28*** | 101.93 | 73.95 | ***94.37*** | 106.75 | 83.37 | ***80.81*** | 102.10 | 64.08 | ***86.03*** | 96.92 | 71.84 | ***78.44*** | 95.24 | 58.98 |
| FEV1 m12 | ***102.55*** | 108.55 | 88.23 | ***83.44*** | 107.65 | 76.71 | ***94.75*** | 105.36 | 85.07 | ***98.65*** | 112.77 | 66.61 | ***88.15*** | 96.60 | 66.53 | ***81.30*** | 94.98 | 67.08 |
| FEV1/FVC w6 | ***103.87*** | 106.94 | 100.85 | ***107.1*** | 110.73 | 97.20 | ***104.35*** | 108.24 | 100.48 | ***101.59*** | 107.26 | 98.29 | ***110.4*** | 115.17 | 104.95 | | | |
| FEV1/FVC m3 | ***102.61*** | 106.39 | 98.04 | ***101.50*** | 107.09 | 94.97 | ***104.00*** | 106.98 | 99.24 | ***105.77*** | 114.48 | 99.53 | ***104.76*** | 112.13 | 99.49 | ***117.10*** | 120.11 | 106.87 |
| FEV1/FVC m6 | ***100.66*** | 103.71 | 97.75 | ***98.76*** | 107.68 | 90.52 | ***102.38*** | 107.75 | 99.33 | ***102.55*** | 120.82 | 99.23 | ***104.8*** | 109.03 | 101.42 | ***108.10*** | 112.04 | 102.61 |
| FEV1/FVC m12 | ***95.62*** | 100.35 | 93.66 | ***96.86*** | 107.99 | 92.01 | ***101.59*** | 106.3 | 98.75 | ***101.98*** | 119.40 | 98.78 | ***104.82*** | 109.63 | 90.81 | ***112.36*** | 118.59 | 97.69 |
| FVC w6 | ***91.91*** | 99.33 | 85.79 | ***81.21*** | 91.13 | 75.50 | ***80.71*** | 90.15 | 68.57 | ***68.50*** | 84.46 | 52.14 | ***61.83*** | 74.64 | 48.14 | | | |
| FVC m3 | ***94.01*** | 101.99 | 87.83 | ***89.35*** | 99.75 | 82.87 | ***88.50*** | 99.32 | 81.69 | ***69.23*** | 92.32 | 50.51 | ***78.38*** | 88.97 | 61.99 | ***50.34*** | 78.60 | 40.97 |
| FVC m6 | ***95.91*** | 104.88 | 92.31 | ***89.61*** | 97.19 | 77.95 | ***93.59*** | 105.07 | 80.24 | ***68.66*** | 101.32 | 60.60 | ***80.23*** | 90.74 | 68.74 | ***75.08*** | 87.88 | 54.93 |
| FVC m12 | ***102.69*** | 109.54 | 87.5 | ***92.50*** | 98.77 | 80.93 | ***93.38*** | 98.20 | 84.31 | ***89.37*** | 107.62 | 61.15 | ***87.29*** | 89.43 | 66.72 | ***79.72*** | 88.26 | 63.25 |
| TLC w6 | ***109.27*** | 124.08 | 99.98 | ***87.78*** | 107.43 | 81.50 | ***87.27*** | 95.93 | 78.63 | ***82.11*** | 102.37 | 60.63 | ***70.92*** | 80.59 | 59.22 | | | |
| TLC m3 | ***103.45*** | 114.53 | 92.46 | ***102.15*** | 113.84 | 92.21 | ***94.88*** | 104.09 | 86.56 | ***86.81*** | 96.46 | 75.07 | ***76.69*** | 93.11 | 71.40 | ***76.01*** | 86.83 | 59.24 |
| TLC m6 | ***112.75*** | 123.58 | 98.99 | ***105.66*** | 117.94 | 89.16 | ***98.44*** | 103.98 | 88.37 | ***75.37*** | 107.34 | 70.67 | ***84.87*** | 92.49 | 74.10 | ***83.53*** | 92.78 | 70.70 |
| TLC m12 | ***112.33*** | 123.32 | 94.085 | ***107.23*** | 108.14 | 104.79 | ***92.85*** | 103.53 | 80.80 | ***87.53*** | 110.67 | 68.95 | ***84.60*** | 95.91 | 70.49 | ***85.08*** | 90.89 | 82.37 |

**Table s1**: Median (bold) (IQR) of patients of different disease severity groups during 6 week, 3, 6 and 12 month follow up. First column in each group shows the median, second the upper IQR limit, third row the lower IQR limit. Abbreviations: w6 – week 6; m3 – month 3; m6 – month 6; m12 – month 12

| **CT-Score** | ***r*** | ***$R^2$*** | ***p*** |
|---|---|---|---|
| **TLC** | -0.44 | 0.19 | <0.0001 |
| **FVC** | -0.30 | 0.089 | 0.0056 |
| **$D_{LCO}$** | -0.30 | 0.090 | 0.0065 |
| **$K_{CO}$** | -0.13 | 0.016 | 0.2440 |

**Table s2:** Pearson correlation coefficient for acute COVID-19 pulmonary involvement as determined by CT-chest score and pulmonary function test parameter during convalescence.

| |
|---|
| Fatigue/exhaustion/excessive tiredness |
| Loss of appetite |
| Apraxia |
| Ataxia |
| Abdominal pain |
| Disturbance of consciousness/confusion/disorientation |
| Conjunctivitis |
| Bleeding |
| Chest pain |
| Diarrhoea |
| Vomiting |
| Facial paresis |
| Fever |
| Gait disorder |
| Memory impairment |
| Sensory disturbance |
| Joint pain |
| Sore throat |
| Skin changes |
| Cough |
| Wheezing |
| Headache |
| Seizures |
| Shortness of breath |
| Paralysis/paresis |
| Runny nose |
| Swelling of the lymph nodes |
| Muscle reflexes (conspicuously reduced/increased) |
| Muscle pain |
| Neglect |
| Neuralgia |
| Sneezing |
| Earache |
| Pyramidal tract signs (positive) |
| Swallowing disorders |
| Dizziness |
| Impaired vision |
| Speech disorders (aphasia) |
| Speech disorders (dysarthria) |
| Disturbances in eye movements |
| Nausea |
| Altered sense of smell or taste |
| Stuffy nose |
| Other symptoms |

**Table s3**: Symptom List of 43 items was assessed at every follow-up study visit
